# Artificial Intelligence–Enabled CMR Tissue Characterization Predicts Reverse Remodeling and Clinical Outcomes in Non-Ischemic Dilated Cardiomyopathy

**DOI:** 10.64898/2025.12.05.25341733

**Authors:** Hyun-Jin Kim, Su A Noh, Kyeong Jin Park, Pan Ki Kim, Minjung Bak, Jiesuck Park, Hong-Mi Choi, Yeonyee E. Yoon, Goo-Yeong Cho, Byoung Wook Choi, Eun Ju Chun, In-Chang Hwang

## Abstract

**Aims:** Diffuse myocardial fibrosis contributes to adverse remodeling and heart failure progression in non-ischemic dilated cardiomyopathy (NIDCM). Quantitative cardiac magnetic resonance (CMR) tissue mapping may improve risk stratification, but manual post-processing limits clinical application. This study aimed to evaluate the prognostic and functional significance of artificial intelligence (AI)–assisted automated quantification of native T1 and extracellular volume fraction (ECV) from baseline CMR in patients with NIDCM.

**Methods and Results:** A total of 347 consecutive patients with NIDCM who underwent baseline CMR at two university-affiliated hospitals between 2018 and 2023 were retrospectively analyzed. An AI algorithm automatically quantified whole-myocardial native T1 and ECV. The primary endpoint was a composite of cardiovascular death or hospitalization for heart failure (HHF). Left-ventricular reverse remodeling (LVRR) was assessed using serial echocardiography. Prognostic performance was assessed using Cox regression and time-dependent receiver-operating characteristic analysis. During a median follow-up of 37.9 months (IQR: 18.1–70.1), 119 patients (34.3%) experienced CV death or HHF. Elevated ECV (≥30%) independently predicted adverse outcomes (adjusted hazard ratio: 3.09; 95% CI: 1.87–5.12; p <0.001), whereas native T1 (≥1325 ms) showed weaker predictive ability. Patients with lower ECV had significantly greater LV functional improvement, while those with higher ECV demonstrated limited LVRR. Individuals with both elevated ECV and native T1 had the lowest composite LVRR rate (52.3%).

**Conclusions:** AI-assisted automated ECV quantification predicts both LVRR and adverse clinical outcomes in NIDCM. The integration of automated CMR tissue mapping into clinical workflows may enhance precision in risk stratification and guide individualized management.

## Introduction

Non-ischemic dilated cardiomyopathy (NIDCM) is characterized by progressive left ventricular (LV) dilation and systolic dysfunction, which leads to adverse remodeling, hospitalization for heart failure (HHF), and premature cardiovascular (CV) death despite advances in medical therapy.(1) Accurate assessment of myocardial tissue is essential for identifying patients at risk of disease progression and for informing individualized management strategies. Conventional imaging metrics, such as LV ejection fraction (LVEF) and chamber size, are insufficient for detecting early or diffuse myocardial fibrosis, a critical determinant of ventricular remodeling and clinical outcomes in NIDCM.(2)

Cardiac magnetic resonance (CMR) imaging with T1 mapping allows for noninvasive quantification of diffuse myocardial fibrosis.(3–5) Native T1 and extracellular volume fraction (ECV) measurements evaluate distinct components of myocardial tissue, specifically the cellular and interstitial compartments.(6) Increased ECV and native T1 values are associated with adverse outcomes and limited LV functional recovery in dilated cardiomyopathy (DCM).(7) However, most studies utilize manual or semi-automated analyses, which are time-intensive, operator-dependent, and difficult to implement in routine clinical practice.(7) Recent developments in artificial intelligence (AI) facilitate automated and reproducible segmentation of the myocardium and tissue mapping.(8–11) However, clinical evidence regarding the utility of these AI-derived measurements in NIDCM is currently limited.

This study examines the clinical value of automated native T1 and ECV measurements in patients with NIDCM. The association between these automated parameters and LV reverse remodeling (LVRR), as well as their prognostic significance for predicting CV death or HHF, was evaluated. It is hypothesized that AI-assisted T1 mapping, particularly automated ECV quantification, can predict LV functional recovery and long-term outcomes, providing a practical and objective tool for risk stratification in NIDCM.

## Method

### Study Population

This retrospective study included 347 consecutive patients diagnosed with NIDCM who underwent baseline CMR imaging between January 2018 and December 2023 at two tertiary hospitals: Seoul National University Bundang Hospital (n = 312) and Hanyang University Guri Hospital (n = 35). NIDCM was defined as LV dilation with systolic dysfunction (LVEF < 50%) in the absence of significant coronary artery disease (CAD). Patients with the following conditions were excluded: (1) significant CAD (≥50% luminal stenosis or prior myocardial infarction), (2) hypertensive heart disease, (3) tachycardia-induced or valvular cardiomyopathy, (4) arrhythmogenic right ventricular cardiomyopathy, (5) LV non-compaction, (6) infiltrative or inflammatory myocardial disease (sarcoidosis, amyloidosis, and myocarditis), (7) chemotherapy- or alcohol-induced cardiomyopathy, or (8) burn-out hypertrophic cardiomyopathy. Patients with an LVEF ≥50% or infarct pattern of late gadolinium enhancement (LGE) on CMR were also excluded.

This study was approved by the Institutional Review Boards of both participating institutions (B-2406-905-104, GURI 2024-06-004), and the requirement for written informed consent was waived owing to the retrospective nature of the study and the use of anonymized data.

### Data Collection

Demographic, clinical, and laboratory data were collected from electronic medical records at the time of baseline CMR. Echocardiographic and CMR datasets were systematically reviewed and analyzed. Transthoracic echocardiography (TTE) was performed using commercially available ultrasound systems, following the current guidelines of the European Association of Cardiovascular Imaging.(12) Measured parameters included LV end-diastolic diameter (LVEDD), left ventricular end-systolic diameter (LVESD), LV end-diastolic volume (LVEDV), LV end-systolic volume (LVESV), LVEF, tricuspid regurgitation maximum velocity (TR Vmax), pulmonary artery systolic pressure (PASP), and LV global longitudinal strain (LVGLS). Follow-up echocardiography was used to quantify changes in LV volume and systolic function over time.

CMR was performed on 3.0-T imager (Ingenia CX, Philips Healthcare, Best, Netherlands) with a 16-channel phased array coil. Cine images using a steady-state free-precession sequence quantified LV and right ventricular (RV) volumes, mass, and ejection fraction. LGE imaging was obtained 10 minutes after intravenous administration of 0.15 mmol/kg gadobutrol using a phase-sensitive inversion-recovery sequence. Native T1-mapping images were acquired using a modified look-locker inversion-recovery 5(3)3 (MOLLI) sequence in three short-axis planes (apical, mid, and base of the LV). Post-contrast T1 mapping images were acquired 15 min after contrast administration along the same three short-axes of LV images used for T1 with a 4(1)3(1)2 sequence. ECV was calculated as ECV = (1–hematocrit) × [△R1_myocardium_]/[△R1_blood_], with hematocrit measured at the time of CMR imaging, where ΔR1 is the change in relaxivity and determined using the following equation:

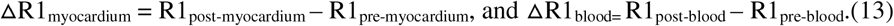

Quantitative tissue parameters were analyzed using an AI-based segmentation and mapping platform (Myomics, Seoul, Republic of Korea). Detailed information, including development and validation, on the AI-algorithm is provided in previous studies.(9–11) The algorithm automatically delineated endocardial and epicardial contours on basal, mid, and apical short-axis slices to compute native T1, post-T1, and ECV values for the entire LV myocardium and mid-septal region. Quality control included visual inspection and manual correction by two experienced CMR specialists who were blinded to clinical outcomes. LGE extent was quantified using signal-intensity thresholds (5 standards deviation (SD) and 6 SD above remote myocardium), and LGE mass and LGE percentage were calculated.

The LVRR was defined as the presence of at least one of the following criteria: an increase in LVEF ≥15 percentage points from baseline; recovery of LVEF to ≥50%; reduction in LVEDV ≥30%, or reduction in LVESV ≥30%.(11) Data of follow-up echocardiography was obtained from the exams performed at an interval of 6–18 months from baseline, and the occurrence of LVRR was determined using the echocardiographic measurements closest to 12 months from baseline, considering that the LVRR typically occurs within 12 months of treatment.(14, 15) Patients without follow-up imaging were excluded from analyses requiring remodeling assessment.

### Clinical Outcomes

The primary endpoint was a composite of CV death or HHF during follow-up. Clinical events were adjudicated through a review of medical records. Follow-up duration was calculated from the date of baseline CMR to the first occurrence of an event or the last known clinical visit.

### Statistical Analysis

Continuous variables are presented as mean±SD for normally distributed data or median with inter-quartile range (25th–75th percentile) for skewed data, and compared using Welch’s *t*-test and Kruskal–Wallis test. Categorical variables are expressed as counts (%) and compared with the Chi-square or Fisher’s exact test. To identify prognostic factors, univariable and multivariable Cox proportional hazards analyses were conducted to evaluate the clinical outcome. Variables with *p* <0.10 in univariable analyses were included in the multivariable model, and the final model was selected using the Akaike Information Criterion (AIC) to determine the optimal combination of predictors. The proportional hazards assumption was verified using Schoenfeld residuals. The predictive performance of continuous imaging biomarkers (ECV and native T1) was assessed using time-dependent receiver operating characteristic (ROC) analysis, which provided optimal cutoff values (30% for ECV and 1325 ms for native T1). Kaplan–Meier survival curves were constructed to visualize event-free survival according to these thresholds and compared using the log-rank test.

All analyses were conducted using Python version 3.11.5 (Python Software Foundation, Wilmington, DE, USA) and R version 4.3.2 (R Foundation for Statistical Computing, Vienna, Austria).

## Results

### Baseline Characteristics

Baseline clinical and laboratory characteristics are presented in **Table 1**. The mean age of study population was 56.2 years (SD 16.1), and 67% were male. Patients who experienced CV death or hospitalization for HHF (n = 119) were older, had a lower body mass index, and had lower blood pressure. Comorbidities such as hypertension and chronic obstructive pulmonary disease (COPD) were more prevalent among patients with adverse outcomes, whereas diabetes mellitus and dyslipidemia showed no significant difference between groups. The use of sodium–glucose cotransporter-2 (SGLT2) inhibitors was significantly lower in the event group, while the other guideline-directed mediations for HF were similar between groups. In laboratory findings, patients with CV death or HHF had significantly lower hemoglobin levels, reduced glomerular filtration rate, and higher B-type natriuretic peptide concentrations, whereas other laboratory parameters showed no significant differences.

**Table 1.**
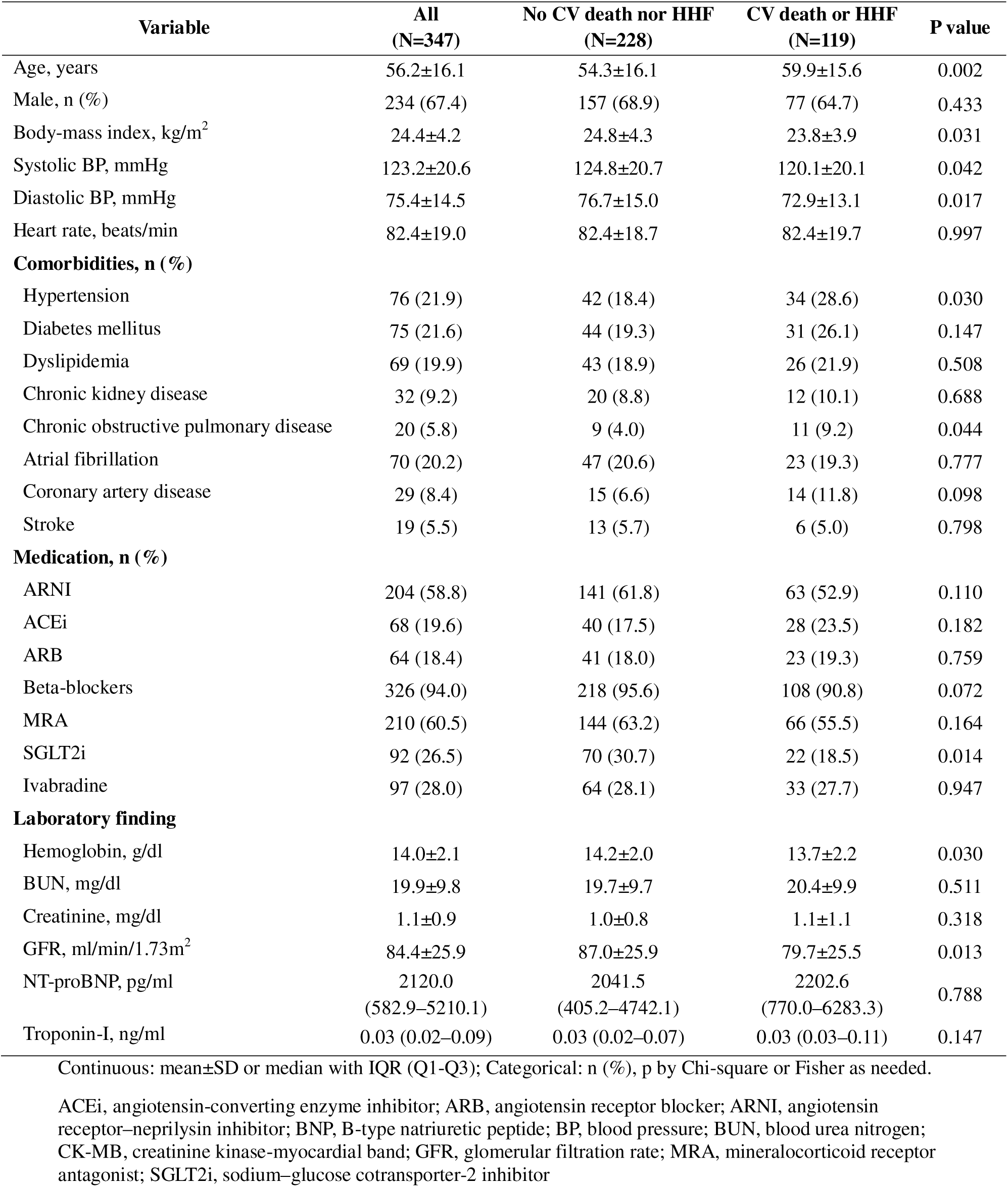
Baseline characteristics.

**Table 2** summarizes the imaging characteristics. CMR-derived indices of LV systolic function and chamber dimensions did not significantly differ between groups. Similarly, RV parameters and LGE burden did not differ significantly. In contrast, CMR tissue characterization metrics demonstrated marked differences. The patients who experienced CV death or HHF had significantly higher ECV and native T1 values, as well as prolonged T2 relaxation times. Echocardiographic parameters revealed no significant differences in LV size, LV systolic function, and RV parameters between groups. LVGLS also did not differ significantly.

**Table 2.** CMR and echocardiographic characteristics.

| Variable | All<br>(N=347) | No CV death nor HHF<br>(N=228) | CV death or HHF<br>(N=119) | P value |
| --- | --- | --- | --- | --- |
| **CMR** |  |  |  |  |
| LVEF, % | 29.6±11.7 | 30.3±12.2 | 28.1±10.7 | 0.075 |
| LVEDV, ml | 253.5±75.9 | 251.3±74.1 | 257.8±79.2 | 0.461 |
| LVESV, ml | 183.4±74.1 | 180.2±74.2 | 189.5±73.7 | 0.269 |
| LV mass, g | 207.8±66.1 | 207.6±67.2 | 208.2±64.1 | 0.931 |
| RVEF, % | 36.8±13.8 | 37.3±13.3 | 35.8±14.7 | 0.348 |
| RVEDV, ml | 147.1±51.7 | 148.4±50.9 | 144.7±53.3 | 0.535 |
| RVESV, ml | 96.1±47.8 | 95.9±46.5 | 96.3±50.4 | 0.948 |
| LGE presence, n (%) | 195 (56.2) | 122 (53.5) | 73 (61.3) | 0.163 |
| LGE mass, g | 106.1±32.9 | 106.9±34.3 | 104.5±30.1 | 0.493 |
| LGE percent 5SD | 2.9±5.6 | 2.6±5.0 | 3.5±6.5 | 0.215 |
| LGE percent 6SD | 2.1±4.5 | 1.8±3.9 | 2.5±5.5 | 0.216 |
| Whole-myocardial Native T1 | 1329.4±60.6 | 1321.8±61.4 | 1346.9±55.4 | 0.001 |
| Mid-Septal Native T1 | 1339.3±64.4 | 1332.6±66.3 | 1354.7±57.2 | 0.007 |
| Whole-myocardial Post_T1 | 464.9±74.4 | 466.5±69.3 | 461.0±85.7 | 0.621 |
| Mid-Septal Post_T1 | 456.8±76.6 | 457.3±70.7 | 455.8±89.7 | 0.897 |
| Whole-myocardial ECV | 27.5±5.8 | 26.8±5.1 | 29.2±6.9 | 0.008 |
| Mid-Septal ECV | 28.5±6.3 | 27.9±5.6 | 29.9±7.5 | 0.035 |
| Whole-myocardial T2 | 51.2±5.1 | 50.6±4.9 | 52.9±5.3 | 0.004 |
| Mid-Septal T2 | 51.1±5.7 | 50.3±4.9 | 53.3±6.9 | 0.003 |
| **Echocardiography** |  |  |  |  |
| LVEDD, mm | 61.1±6.9 | 61.1±6.7 | 61.2±7.3 | 0.825 |
| LVESD, mm | 51.7±8.5 | 51.8±8.2 | 51.5±9.0 | 0.766 |
| LVEDV, ml | 168.4±54.3 | 170.4±54.8 | 164.7±53.4 | 0.356 |
| LVESV, ml | 120.6±50.9 | 122.5±51.6 | 117.0±49.4 | 0.339 |
| LVEF, % | 29.2±10.3 | 29.5±10.8 | 28.6±9.3 | 0.433 |
| TR Vmax, m/s | 2.6±0.5 | 2.5±0.5 | 2.6±0.6 | 0.148 |
| PASP, mmHg | 33.6±13.2 | 32.9±12.2 | 35.0±14.8 | 0.178 |
| LVGLS, % | 10.9±4.4 | 11.1±4.5 | 10.4±4.1 | 0.195 |

ECV, extracellular volume fraction; LGE, late gadolinium enhancement; LVEDD, left ventricular end-diastolic diameter; LVEDV, left ventricular end-diastolic volume; LVEF, left ventricular ejection fraction; LVESD, left ventricular end-systolic diameter; LVESV, left ventricular end-systolic volume; LVGLS, left ventricular global longitudinal strain; LV mass, left ventricular mass; PASP, pulmonary artery systolic pressure; RVEF, right ventricular ejection fraction; RVEDV, right ventricular end-diastolic volume; RVESV, right ventricular end-systolic volume; TR Vmax, tricuspid regurgitation maximum velocity.

### Clinical Outcomes

During a median follow-up of 37.9 months (interquartile range: 18.1–70.1), a total of 119 patients (34.3%) experienced the composite clinical endpoint. **Figure 1** presents the time-dependent ROC analysis of baseline CMR tissue parameters for predicting adverse clinical outcomes. Both the whole-myocardial ECV and the whole-myocardial native T1 demonstrated significant discriminatory capacity for the composite clinical outcome, with optimal cutoff values of 30% for ECV and 1325 ms for native T1. The corresponding areas under the curve (AUC) were 0.67 (sensitivity 0.46, specificity 0.87) and 0.63 (sensitivity 0.71, specificity 0.52), respectively. **Figures 2A** and **2B** depict Kaplan–Meier survival curves stratified by these cutoff thresholds. Patients with ECV ≥30% or native T1 ≥1325 ms experienced significantly higher cumulative rates of CV death or HHF compared with those below the thresholds (both log-rank p <0.001),

**Figure 1.**
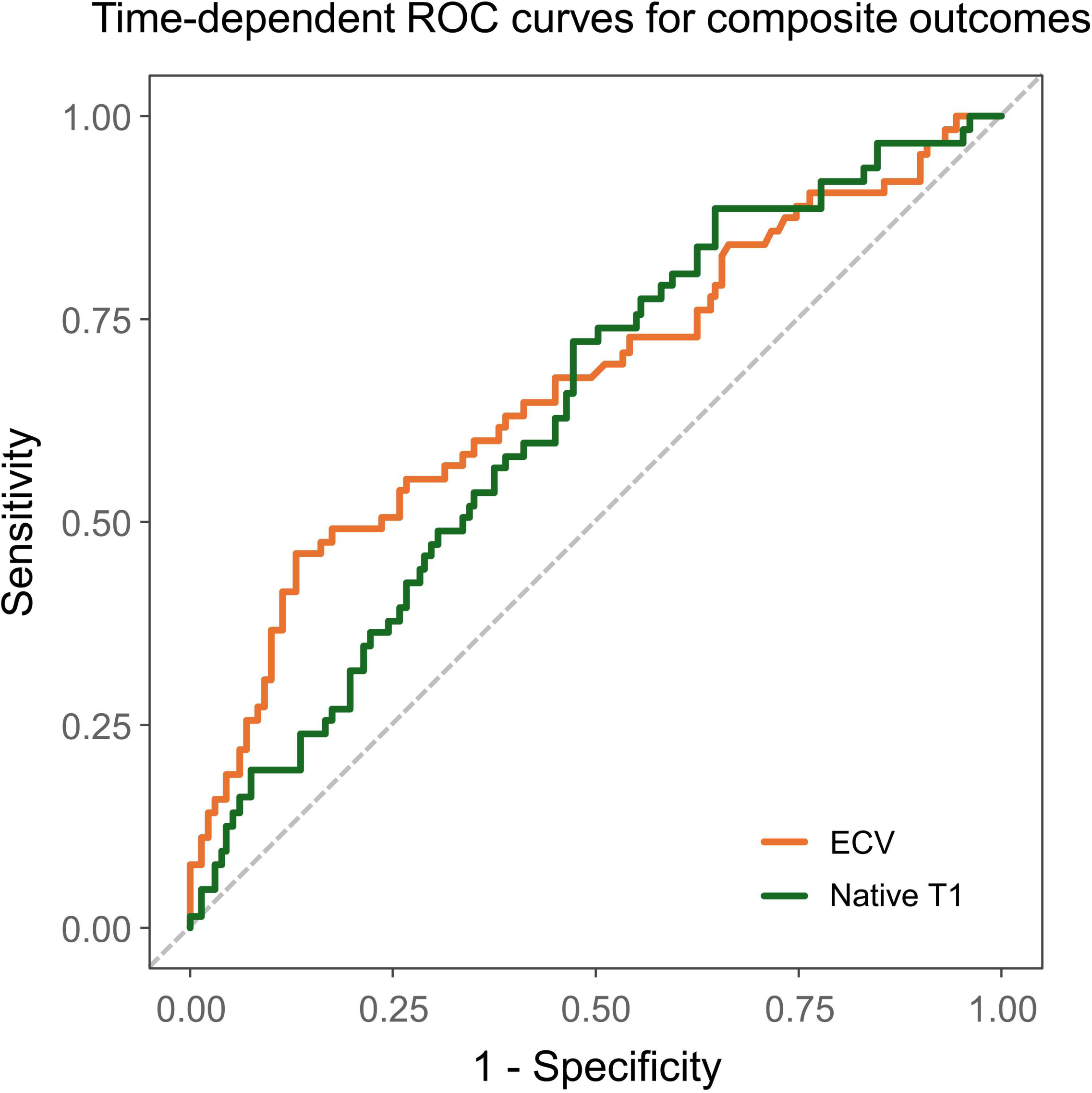
ROC curves for clinical outcome. Time-dependent receiver operating characteristic (ROC) curves for predicting cardiovascular death or heart failure hospitalization based on cardiac magnetic resonance T1-mapping parameters. Whole-myocardial extracellular volume fraction (ECV) and native T1 demonstrated significant prognostic accuracy, with optimal cutoff values of 30% and 1325 ms, respectively.

**Figure 2.**
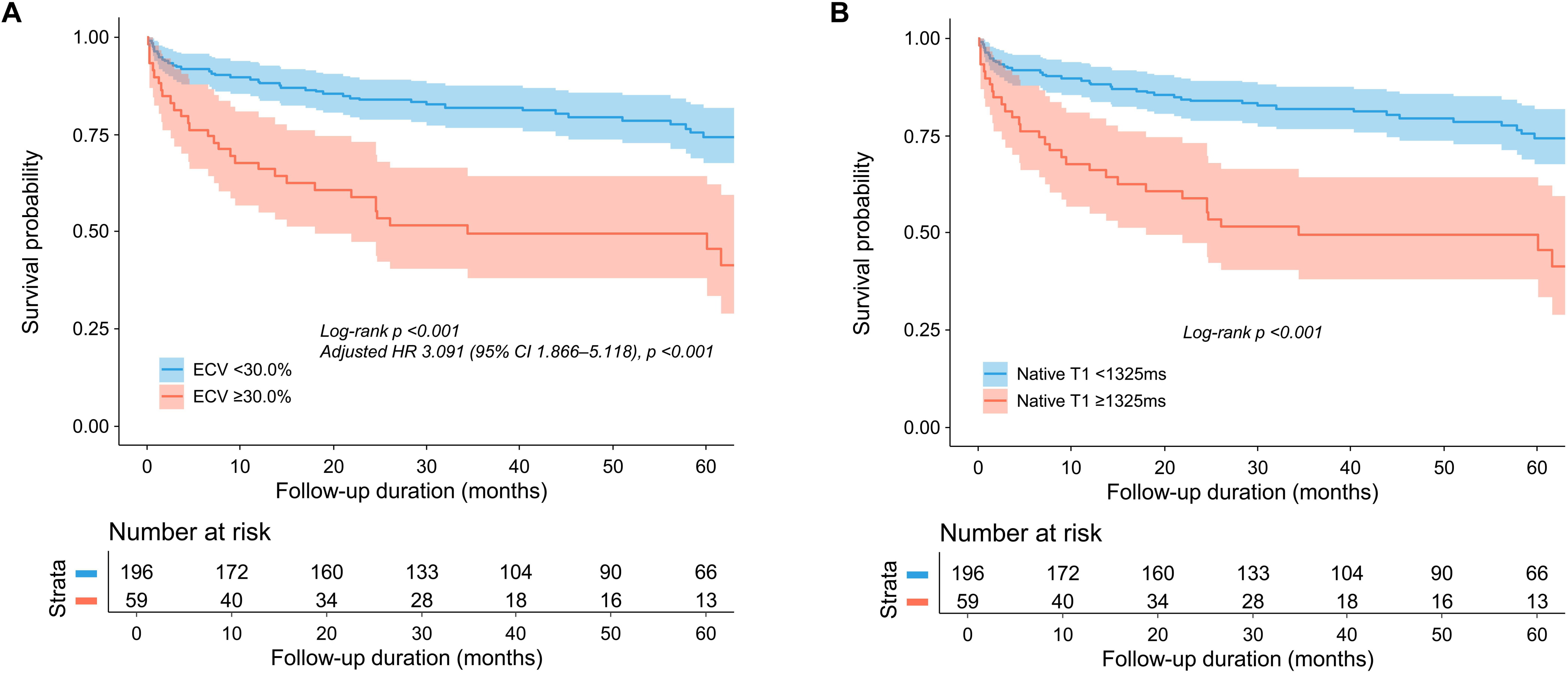
Kaplan-Meier survival curve according to T1-mapping parameters. **(A)** Kaplan–Meier survival curves stratified by whole-myocardial ECV ≥30% versus <30%. **(B)** Survival curves according to whole-myocardial native T1 ≥1325 ms versus <1325 ms.

### Left Ventricular Reverse Remodeling According to CMR T1-Mapping Parameters

The association between the achievement of LVRR and the whole-myocardial ECV and whole-myocardial native T1 was assessed. The median interval between baseline and follow-up TTE was 12.1 months (interquartile range: 9.0–14.9 months). **Figure 3A** demonstrates that patients with lower ECV values (<30%) experienced significantly greater improvement in LV systolic and volumetric parameters compared to those with higher ECV values. The low-ECV group exhibited substantially larger reductions in LVEDV and LVESV, both in absolute terms and as percentage change from baseline. Improvement in LVEF was also significantly greater among patients with ECV <30%. Detailed categorical analyses of LVRR components showed consistent findings that lower ECV at baseline was significantly associated with favorable LV structural and functional improvements (**Supplementary Table 1**). In contrast, **Figure 3B** shows that when patients were stratified by native T1 <1325 ms vs ≥1325 ms, there were no significant differences in LVEDV, LVESV, or LVEF changes (all p >0.1). While categorical analysis revealed that the proportion of patients achieving composite LVRR was significantly lower among those with higher native T1 values (≥ 1325 ms) compared to those with lower values, no significant trends were observed for individual detailed remodeling parameters when stratified by native T1 levels (**Supplementary Table 1**). Additionally, patients with both elevated ECV (≥30%) and elevated native T1 (≥1325 ms) had the lowest rate of composite LVRR (52.3%), (**Supplementary Table 2**).

**Figure 3.**
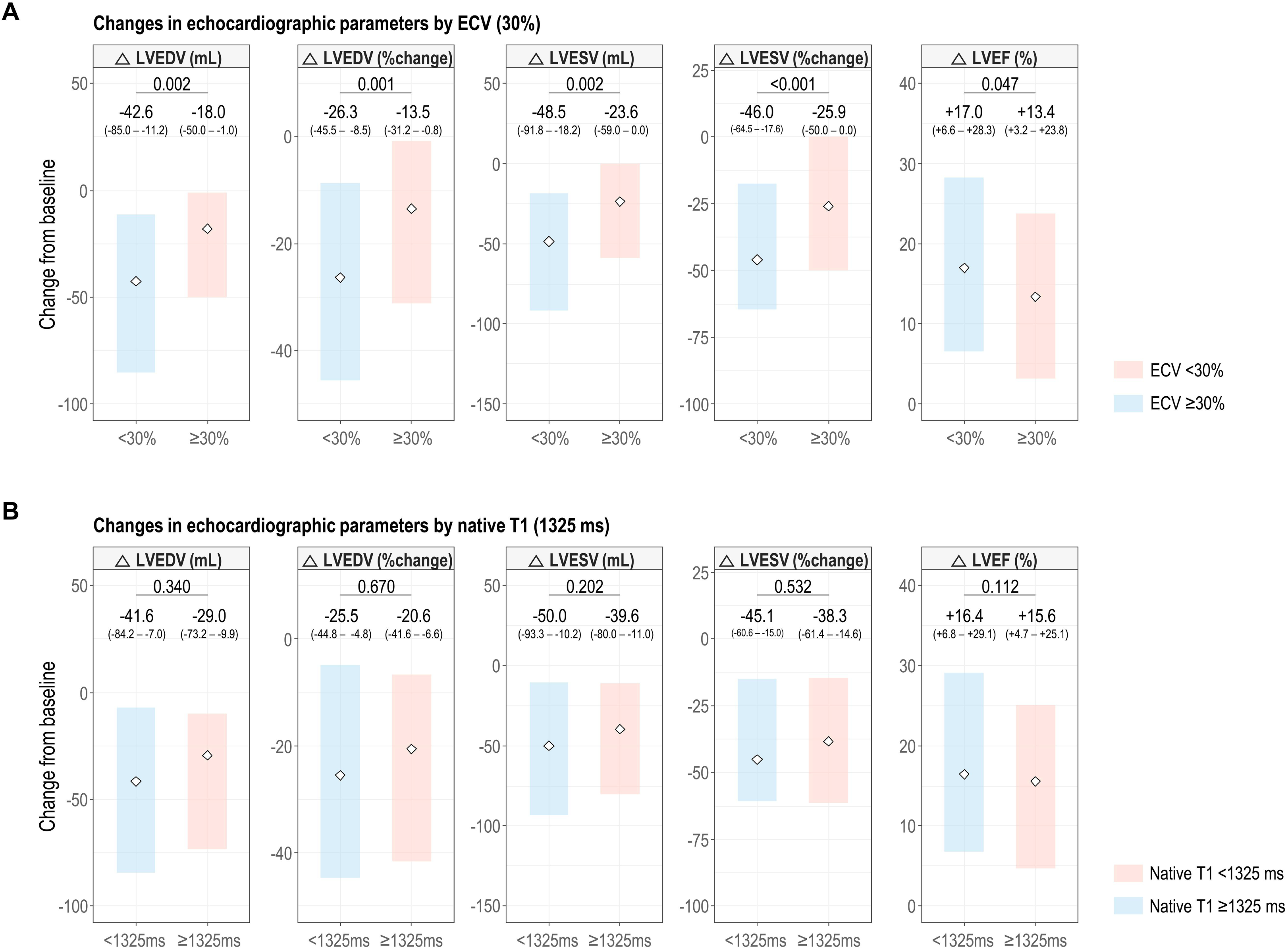
Echocardiographic changes according to ECV and native T1 cutoff values. Changes in LVEDV, LVEDV % change, LVESV, LVESV %, LVEF change were compared between subgroups stratified by whole-myocardial ECV <30% vs. ≥30% (A), and by whole-myocardial native T1 <1325 ms vs ≥1325 ms (B). Error bars represent standard error of the mean; p-values were calculated using Welch’s t-test. ECV, extracellular volume; LV, left-ventricular; LVEDV, left-ventricular end-diastolic volume; LVEF, left-ventricular ejection fraction; LVESV, LV end-systolic volume

### Predictors of Clinical Outcomes

We used Cox proportional hazards analyses of baseline clinical and CMR-derived parameters to identify independent predictors of CV death or HHF (**Table 3**). In the univariable analysis, older age, higher ECV (ECV ≥30%), the absence of LVRR, lower diastolic blood pressure, and the presence of hypertension, CAD, or COPD were each associated with increased risk of the composite endpoint. Among T1-mapping parameters, elevated ECV demonstrated a significant univariable association with adverse events (hazard ratio [HR], 2.92; 95% CI, 1.841–4.631; p <0.001), and the presence of LGE showed a tendency for a higher risk of adverse events (HR 1.427; 95% CI, 0.99–2.07; p = 0.060). The multivariable model, optimized using AIC, identified age, elevated ECV, presence of LGE, failure to achieve composite LVRR, hypertension, and COPD as independent predictors of the outcome. Elevated ECV ≥30% was a significant predictor of clinical outcomes (adjusted HR 2.00, 95% CI 1.32–3.03, p <0.001) after adjusting for age and comorbidities, whereas the presence of LGE was not associated with the clinical outcomes in the multivariable analysis. Hypertension and COPD also increased risk (hypertension: adjusted HR 1.56, 95% CI 1.04–2.34, p = 0.032; COPD: adjusted HR 1.89, 95% CI 1.01–3.55, p = 0.047). The failure to achieve composite LVRR was significantly associated with clinical outcomes (adjusted HR 1.60; 95% CI 1.04–2.45; p = 0.031). The final AIC-selected Cox model showed good discriminative performance (AUC = 0.790) and stability.

**Table 3.** Predictors of clinical outcome.

|  | Univariable analysis |  |  | Multivariate |  |  |
| --- | --- | --- | --- | --- | --- | --- |
|  | Unadjusted HR | 95% CI | P value | Adjusted HR | 95% CI | P value |
| Age | 1.02 | 1.01–1.03 | 0.001 | 1.01 | 1.00–1.03 | 0.006 |
| Whole-myocardial ECV $\geq 30\%$ | 2.92 | 1.84–4.63 | <0.001 | 2.00 | 1.32–3.03 | 0.001 |
| Presence of LGE | 1.43 | 0.99–2.07 | 0.060 | 1.32 | 0.91–1.92 | 0.146 |
| Failure to achieve composite LVRR | 1.61 | 1.04–2.51 | 0.034 | 1.60 | 1.04–2.45 | 0.031 |
| Systolic BP | 0.99 | 0.98–1.00 | 0.052 | - | - | - |
| Diastolic BP | 0.99 | 0.97–0.99 | 0.028 | - | - | - |
| Hypertension | 1.67 | 1.12–2.48 | 0.012 | 1.56 | 1.04–2.34 | 0.032 |
| Diabetes mellitus | 1.42 | 0.94–2.13 | 0.098 | - | - | - |
| Coronary artery disease | 1.90 | 1.09–3.32 | 0.025 | - | - | - |
| Chronic obstructive pulmonary disease | 2.06 | 1.11–3.83 | 0.023 | 1.89 | 1.01–3.55 | 0.047 |

Multivariable model selected by Akaike Information Criterion (AIC) minimization among nine candidate variables (Age, ECV $\geq 30\%$ , composite LVRR, systolic BP, diastolic BP, hypertension, diabetes mellitus, coronary artery disease, chronic obstructive pulmonary disease). For multivariable modeling, all variables with $p < 0.10$ in univariate analyses were entered into an AIC-based stepwise Cox proportional hazards model.

## Discussion

In this study, we evaluated patients with NIDCM who underwent baseline CMR imaging, analyzed using an AI-assisted algorithm to automatically quantify native T1 and ECV (**Graphical Abstract**). The principal findings were as follows: first, AI-derived quantitative CMR T1-mapping parameters, particularly ECV, were strongly associated with cardiovascular outcomes, including CV death and HHF. Second, mechanistic analysis revealed that patients with lower baseline ECV values exhibited a greater likelihood of LV functional recovery and reverse remodeling during follow-up, whereas those with elevated ECV showed persistently impaired LV contractile improvement. Collectively, these results indicate that AI-based automated ECV quantification provides prognostic value and may help predict clinical outcomes and myocardial recovery in NIDCM. Conversely, native T1, while correlated with ECV, appeared less effective in predicting prognosis or the degree of LV functional improvement, suggesting that diffuse extracellular expansion rather than intracellular changes predominantly drives outcome differences in this population.

Histopathologic validation studies have demonstrated that ECV closely corresponds to diffuse interstitial collagen deposition, which disrupts myocardial architecture, impairs contractility, and limits reverse remodeling.(16–18) Puntmann et al. reported that T1-mapping indices (including ECV) independently predicted HHF and mortality even after adjustment for LVEF and LGE burden.(19) Similarly, Vita et al. demonstrated in a cohort of patients with NIDCM that ECV was the strongest and only independent predictor of heart failure outcomes after adjustment for clinical and imaging covariates.(18) Furthermore, Li et al. and Cadour et al. confirmed that elevated ECV remains an independent predictor of mortality and HHF in NIDCM cohort.(7, 20) The present study extends these observations by demonstrating that AI-based automated ECV quantification retains robust prognostic relevance and correlates with subsequent LV functional improvement. In contrast, native T1—while sensitive to both intracellular and extracellular changes—showed weaker prognostic performance. This may be explained by the fact that native T1 represents a composite signal influenced by myocardial water content and inflammation rather than pure extracellular expansion,(21) and mildly increased native T1 may reflect reversible processes such as edema.(22, 23) Taken together, these results reinforce the concept that the extent of extracellular matrix expansion represents a mechanistic substrate of disease progression and resistance to reverse remodeling in NIDCM. They further suggest that quantitative ECV mapping can serve as a non-invasive surrogate for myocardial reparability, enabling identification of patients with preserved versus exhausted reserve and providing insights into disease reversibility.

Our findings highlight the potential clinical utility of AI-assisted CMR tissue mapping as a noninvasive tool for risk stratification and personalized management in DCM. Automated quantification of myocardial ECV enables rapid and reproducible assessment of diffuse interstitial fibrosis, which can identify patients with limited potential for reverse remodeling despite optimized guideline-directed medical therapy.(24) In clinical practice, such patients may benefit from closer surveillance, early consideration of advanced therapies, or timely referral for device-based interventions. Conversely, individuals with low ECV and preserved remodeling capacity may be managed more conservatively, thereby avoiding unnecessary escalation of treatment. The integration of AI-based tissue characterization into standard CMR workflows could thus improve prognostic precision while reducing interobserver variability inherent in manual post-processing.(25)

The automated nature of AI-based T1-mapping also facilitates the analysis of the entire LV myocardium, rather than restricting measurement to the mid-septal region. The mid-septal region-of-interest approach has traditionally been acceptable for assessing myocardial fibrosis on CMR, with advantages in minimizing artifacts from adjacent structures.(26) Given that LGE in NIDCM typically involves the basal to mid-septal myocardium, this approach retains clinical relevance. However, regional variation in ECV values has been reported, with septal ECV tending to be slightly higher than non-septal myocardium.(27) In a study of DCM, septal segments showed the highest ECV values, but only global ECV–not septal ECV–was significantly associated with clinical outcomes such as arrhythmic burden.(28) Similarly, in patients with NIDCM, native T1, ECV, and LGE demonstrated variable prognostic significance across six LV segments; ECV was consistently associated with adverse outcomes across all regions, though the strength of association varied by location.(18) Moreover, the number of segments with abnormal ECV showed a linear relationship with the risk of adverse events. Collectively, these findings suggest that T1-mapping-derived tissue characteristics may have greater clinical relevance when assessed across the entire LV myocardium rather than a single region. In line with these studies, our analysis demonstrated that mid-ECV at mid-septum is higher than the whole myocardial ECV, and that prognostic significance was observed only for whole-myocardial ECV. Thus, AI-assisted T1-mapping enables large-scale, operator-independent quantification of myocardial tissue characteristics–such as ECV, native T1, and LGE–with markedly reduced interpretation time, supporting broader clinical and research applications and facilitating multicenter standardization and serial monitoring of myocardial remodeling over time

This study has several limitations. First, the moderate sample size from two university-affiliated hospitals may limit the generalizability of our findings to broader DCM populations. Second, not all patients had follow-up echocardiography, which could introduce selection bias in reverse remodeling analyses. Third, we did not obtain histological confirmation of myocardial fibrosis, so elevated ECV should be considered a surrogate marker. Finally, changes in treatment during follow-up, including the new heart failure therapies, may have influenced outcomes but were not fully captured in our models.

## Conclusion

In patients with NIDCM, CMR with AI-assisted T1 mapping provided important prognostic information. Elevated ECV was strongly associated with adverse outcomes, including CV death and HHF, and predicted a reduced likelihood of subsequent LVRR. In contrast, native T1 was less effective in forecasting functional recovery or long-term prognosis. These findings suggest that AI-derived automated ECV quantification reflects irreversible interstitial fibrosis and may serve as a robust imaging biomarker for risk stratification and therapeutic decision-making in DCM.

## Data Availability Statement

All data produced in the present study are available upon reasonable request to the authors.

