## Supplementary Table S1 and S2 for "Artificial Intelligence–Enabled CMR Tissue Characterization Predicts Reverse Remodeling and Clinical Outcomes in Non-Ischemic Dilated Cardiomyopathy"

**Running Title: AI-Based T1 mapping CMR for Prognosis in DCM**

Hyun-Jin Kim, MD ^1*^, Su A Noh, MD ^2^*, Kyeong Jin Park, MD ^3^,

Pan Ki Kim, PhD ^4^, Minjung Bak, MD ^2^, Jiesuck Park, MD ^2,5^, Hong-Mi Choi, MD ^2,5^,

Yeonyee E. Yoon, MD, PhD ^2,5^, Goo-Yeong Cho, MD, PhD ^2,5^,

Byoung Wook Choi, MD, PhD ^4,6^, Eun Ju Chun, MD, PhD ^3,7†^, In-Chang Hwang, MD ^2,5†^

^1^ Division of Cardiology, Department of Internal Medicine, Hanyang University College of Medicine, Hanyang University Guri Hospital, Guri, Gyeonggi, Korea

^2^ Cardiovascular Center, Seoul National University Bundang Hospital, Seongnam, Gyeonggi, Korea

^3^ Department of Radiology, Seoul National University Bundang Hospital, Seongnam, Gyeonggi, Korea

^4^ Phantomics, Inc., Seoul, Korea

^5^ Department of Internal Medicine, Seoul National University College of Medicine, Seoul, Korea

^6^ Department of Radiology, Research Institute of Radiological Science, Center for Clinical Imaging Data Science, Yonsei University College of Medicine, Seoul, Korea

^7^ Department of Radiology, Seoul National University College of Medicine, Seoul, Korea

***** These authors contributed equally to this work as co-first authors**.**

**†** These authors contributed equally to this work as co-corresponding authors**.**

**Supplementary Table 1. Association of baseline whole-myocardial ECV and native T1 with LV reverse remodeling**

|  | **ECV ≥ 30%**  **(n = 60)** | **ECV < 30%**  **(n = 196)** | **P value** | **Odds ratio** | **95% CI** | **P value (logistic)** |
| --- | --- | --- | --- | --- | --- | --- |
| Composite LVRR | 29 (54.7) | 128 (71.9) | 0.019 | 0.47 | 0.25–0.89 | 0.020 |
| ≥15% reduction in LVEDV | 24 (45.3) | 115 (64.6) | 0.012 | 0.45 | 0.24–0.84 | 0.013 |
| ≥30% reduction in LVEDV | 14 (26.4) | 79 (44.4) | 0.019 | 0.45 | 0.23–0.89 | 0.021 |
| ≥15% reduction in LVESV | 34 (64.2) | 138 (77.5) | 0.050 | 0.52 | 0.27–1.01 | 0.052 |
| ≥30% reduction in LVESV | 24 (45.3) | 117 (65.7) | 0.007 | 0.43 | 0.23–0.80 | 0.008 |
| ≥10% absolute increase in LVEF | 29 (53.7) | 121 (68.0) | 0.055 | 0.55 | 0.29–1.02 | 0.056 |
| ≥15% absolute increase in LVEF | 25 (46.3) | 101 (56.7) | 0.177 | 0.66 | 0.36–1.21 | 0.179 |
| Recovery of LVEF to >50% | 17 (31.5) | 75 (42.1) | 0.161 | 0.63 | 0.33–1.20 | 0.163 |
| ≥15% reduction in LVEDV and ≥10% increase in LVEF | 20 (37.7) | 96 (53.9) | 0.038 | 0.52 | 0.28–0.97 | 0.040 |
| ≥30% reduction in LVEDV and ≥10% increase in LVEF | 13 (24.5) | 73 (41.0) | 0.029 | 0.47 | 0.23–0.94 | 0.032 |
|  | **Native T1 ≥1325 ms**  **(n = 137)** | **Native T1 <1325ms**  **(n = 124)** | **P value** | **Odds ratio** | **95% CI** | **P value (logistic)** |
| Composite LVRR | 77 (62.1) | 84 (75.0) | 0.034 | 0.55 | 0.31–0.96 | 0.035 |
| ≥15% reduction in LVEDV | 73 (58.9) | 70 (62.5) | 0.569 | 0.86 | 0.51–1.45 | 0.569 |
| ≥30% reduction in LVEDV | 50 (40.3) | 45 (40.2) | 0.982 | 1.01 | 0.60–1.69 | 0.982 |
| ≥15% reduction in LVESV | 92 (74.2) | 84 (75.0) | 0.887 | 0.96 | 0.53–1.72 | 0.887 |
| ≥30% reduction in LVESV | 72 (58.1) | 73 (65.2) | 0.262 | 0.74 | 0.44–1.25 | 0.263 |
| ≥10% absolute increase in LVEF | 77 (61.6) | 77 (68.8) | 0.249 | 0.73 | 0.43–1.25 | 0.250 |
| ≥15% absolute increase in LVEF | 65 (52.0) | 64 (57.1) | 0.427 | 0.81 | 0.49–1.36 | 0.428 |
| Recovery of LVEF to >50% | 44 (35.2) | 49 (43.8) | 0.178 | 0.70 | 0.41–1.18 | 0.179 |
| ≥15% reduction in LVEDV and ≥10% increase in LVEF | 61 (49.2) | 59 (52.7) | 0.593 | 0.87 | 0.52–1.45 | 0.593 |
| ≥30% reduction in LVEDV and ≥10% increase in LVEF | 47 (37.9) | 41 (36.6) | 0.837 | 1.06 | 0.62–1.79 | 0.837 |

CI, confidence interval; ECV, extracellular volume fraction; LV, left ventricle; LVEDV, left ventricular end-diastolic volume; LVEF, left ventricular ejection fraction; LVESV, left ventricular end-systolic volume; LVRR, left ventricular reverse remodeling.

**Supplementary Table 2. Association of baseline ECV and native T1 with LV reverse remodeling**

|  | **Native T1 <1325ms**  **& ECV <30%**  **(n = 110)** | **Native T1 <1325ms**  **& ECV ≥ 30%**  **(n =12)** | **Native T1 ≥1325 ms**  **& ECV <30%**  **(n =86)** | **Native T1 ≥1325 ms**  **& ECV ≥ 30%**  **(n=48)** | **P value** |
| --- | --- | --- | --- | --- | --- |
| Composite LVRR | 76 (76.77) | 7 (63.64) | 52 (65.82) | 22 (52.38) | 0.037 |
| ≥15% reduction in LVEDV | 64 (64.65) | 5 (45.45) | 51 (64.56) | 19 (45.24) | 0.095 |
| ≥30% reduction in LVEDV | 44 (44.44) | 1 (9.09) | 35 (44.30) | 13 (30.95) | 0.065 |
| ≥15% reduction in LVESV | 78 (78.79) | 5 (45.45) | 60 (75.95) | 29 (69.05) | 0.087 |
| ≥30% reduction in LVESV | 68 (68.69) | 4 (36.36) | 49 (62.03) | 20 (47.62) | 0.037 |
| ≥10% absolute increase in LVEF | 69 (69.70) | 7 (63.64) | 52 (65.82) | 22 (51.16) | 0.205 |
| ≥15% absolute increase in LVEF | 57 (57.58) | 7 (63.64) | 44 (55.70) | 18 (41.86) | 0.313 |
| Recovery of LVEF to >50% | 45 (45.45) | 4 (36.36) | 30 (37.97) | 13 (30.23) | 0.372 |
| ≥15% reduction in LVEDV and ≥10% increase in LVEF | 54 (54.55) | 4 (36.36) | 42 (53.16) | 16 (38.10) | 0.228 |
| ≥30% reduction in LVEDV and ≥10% increase in LVEF | 40 (40.40) | 1 (9.09) | 33 (41.77) | 12 (28.57) | 0.102 |

ECV, extracellular volume fraction; LV, left ventricle; LVEDV, left ventricular end-diastolic volume; LVEF, left ventricular ejection fraction; LVESV, left ventricular end-systolic volume; LVRR, left ventricular reverse remodeling
